# Containment of a multi-index B.1.1.7 outbreak on a university campus through a genomically-informed public health response

**DOI:** 10.1101/2022.01.04.22268758

**Authors:** Emily T. Martin, Adam S. Lauring, JoLynn P. Montgomery, Andrew L. Valesano, Marisa C. Eisenberg, Danielle Sheen, Jennifer Nord, Robert D. Ernst, Lindsey Y. Mortenson, Riccardo Valdez, Yashar Niknafs, Darryl Conway, Sami F. Rifat, Natasha Bagdasarian, Sarah Lyon-Callo, Jim Collins, Heather Blankenship, Marty Soehnlen, Juan Marquez

**Affiliations:** Department of Epidemiology, University of Michigan School of Public Health; Department of Internal Medicine, University of Michigan School of Medicine; EpiStudies, LLC; Environmental Health & Safety, University of Michigan; University Health Service, University of Michigan; Department of Pathology, University of Michigan Medical School; LynxDx, Inc; Athletics, University of Michigan; Athletic Medicine, University of Michigan; Michigan Department of Health and Human Services; Washtenaw County Health Department

## Abstract

The first cluster of SARS-CoV-2 cases with lineage B.1.1.7 in the state of Michigan was identified through intensive university-led surveillance sampling and targeted sequencing. A collaborative investigation and response was conducted by the local and state health departments, and the campus and athletic medicine COVID-19 response teams, using S-gene target failure screening and rapid genomic sequencing to inform containment strategies. A total of 50 cases of B.1.1.7-lineage SARS-CoV-2 were identified in this outbreak, which was due to three coincident introductions of B.1.1.7-lineage SARS-CoV-2, all of which were genetically distinct from lineages which later circulated in the broader community. This investigation demonstrates the successful implementation of a genomically-informed outbreak response which can be extended to university campuses and other settings at high risk for rapid emergence of new variants.

## Introduction

The continued emergence of SARS-CoV-2 variants has brought much attention to the need for genomic surveillance at both global and local scales. Despite rapid advances in the scale and availability of SARS-CoV-2 sequencing, challenges remain in determining how best to make sequencing data actionable at an individual level as part of the case investigation and contact tracing process. The emergence of the B.1.1.7 variant, first identified in the United Kingdom in late 2020, was an early demonstration of the benefit and challenges of interpreting SARS-CoV-2 genomic and PCR data (based on S-gene target failure or SGTF patterns) within actively evolving public health practice. The increased transmissibility(1,2) of B.1.1.7 required a reconsideration of mitigation strategies(3) compared to those used with earlier lineages. This is a challenge that continues to persist with the emergence of highly-transmissible Delta and now Omicron variants.

The identification of the B.1.1.7 variant and initial characterization of its epidemiology in the United Kingdom occurred after it was already present throughout the region, and after widespread community transmission was established. For this reason, there have been few opportunities to describe new B.1.1.7 introductions into a community and the subsequent spread of the variant. University communities are unique environments from which to detect and mitigate the spread of variants of concern. Students have larger networks of contacts compared to the non-university community, and congregate living is more common, whether in residence halls or in large houses. At the start of each semester or during academic breaks(4,5), the frequency of travel to the local campus area from both domestic and international locations facilitates new introductions of SARS-CoV-2 viruses from other regions. Many universities have implemented asymptomatic testing programs at various scales. These programs offer a level of testing that extends beyond what is available to the general public. This increased testing enables earlier identification of introductions of virus and, when coupled with genomic surveillance, earlier identification of variants of concern. While important for informing initiatives to reduce transmission on campus, these efforts also give a more detailed picture of virus introduction and spread than what is possible to observe outside of a university campus.

On January 16, 2021, the first cluster of SARS-CoV-2 cases with lineage B.1.1.7 (Alpha) in the state of Michigan was identified through intensive university-led surveillance sampling followed by targeted sequencing by the Michigan Department of Health and Human Services (MDHHS) Bureau of Laboratories. A collaborative investigation and response was conducted jointly by the local and state health departments, and the campus and athletic medicine COVID-19 response teams. This investigation paired genomic sequencing with enhanced case investigation and contact tracing to identify and contain the outbreak. Through coupling data from a university student testing program with rapid turnaround genomic surveillance, we were able to trace the emergence, spread and containment of the B.1.1.7 variant in a university campus community. Here we describe the genomic epidemiology of the variant as well as the testing and mitigation efforts that combined to ensure early identification and rapid containment of the outbreak.

## Methods

### Initial Identification

The initial B.1.1.7 isolate was identified in an individual who was participating in an athletics-wide testing program for all student athletes; this included return to campus reverse transcription-polymerase chain reaction (RT-PCR) testing, daily conference-mandated rapid antigen testing with RT-PCR confirmation of positive results for in-season athletes, and once to thrice weekly (depending on sport) RT-PCR for out-of-season athletes. This initial specimen was sent to MDHHS for whole genome sequencing based on recent travel to the United Kingdom.

### Testing Protocol

An enhanced testing and genomic sequencing protocol was initiated to identify the lineage of all university-associated cases upon identification of the first B.1.1.7 case. All students were required to complete a daily app-based symptom screen, and symptomatic students were referred to university health clinics for symptomatic testing by RT-PCR. These tests were performed either at the Michigan Medicine Clinical Microbiology or Molecular Diagnostic Laboratories or the University (of Michigan) Health Services laboratory. During the outbreak investigation, the ThermoFisher TaqPath assay was used as a preliminary screen while awaiting sequencing results, either as the initial RT-PCR method, or to screen positives identified by other testing platforms. Observation of an S Gene Target Failure (SGTF) on this assay was considered a presumptive detection of a B.1.1.7 variant infection. Individuals with an SGTF result and an epidemiologic linkage to a known B.1.1.7 case were classified as probable B.1.1.7 cases for the purposes of the outbreak investigation.

### Whole Genome Sequencing

Specimens from cases identified in January and February were analyzed by whole genome sequencing by MDHHS Bureau of Laboratories or by a laboratory at the University. In addition, all available positive specimens from non-University affiliated individuals seeking care at Michigan Medicine were also processed for sequencing. Residual nasopharyngeal and saliva specimens from symptomatic and asymptomatic individuals who tested positive for SARS-CoV-2 by RT-PCR were obtained from the Michigan Medicine Clinical Microbiology Laboratory, University Health Services, and LynxDx (Ann Arbor, MI). Specimens from individuals living in congregate settings (residence halls, or fraternity, sorority or cooperative houses), from individuals with recent travel, or from student athletes were prioritized for sequencing on the Nanopore platform, which allowed for more rapid identification of B.1.1.7 cases than with the Illumina platform (Supplementary Figure 1). Specimens were processed using the Artic Network v3 primer pools and protocol(6,7).

### Phylogenetic methods

To generate a phylogenetic tree, we aligned consensus genomes with MUSCLE 3.8.31 and masked positions that are known to commonly exhibit homoplasies or sequencing errors(8). We generated a maximum likelihood phylogeny with IQ-TREE, using a GTR model and 1000 ultrafast bootstrap replicates(9,10). Evolutionary lineages (Pango lineages) were assigned with PANGOLIN(11). Pangolin lineage assignment with >90% genome coverage was considered confirmatory laboratory evidence of B.1.1.7. Sequencing results with less than 90% coverage but with greater than 50% of hallmark lineage mutations detected and over 50% of genome coverage were considered presumptive laboratory evidence of B.1.1.7. Individuals with confirmatory sequence-based laboratory evidence of B.1.1.7 were classified as confirmed B.1.1.7 cases for the purposes of the outbreak investigation.

### Estimate of R_t_

The time varying effective reproduction number, R_t_, was estimated based on the B.1.1.7 case time series data, assuming a mean serial interval of 5.68 and serial interval standard deviation (SD) of 4.77 (based on estimates from Reed et al(12)). R_t_ was estimated using the EpiEstim package developed by Cori et al.(13) using a 7-day sliding window. As a sensitivity analysis, we also tested a wider range of serial interval distributions ranging over ±2 from the estimated serial interval mean and SD (i.e. serial interval mean range of 3.68–7.68 and SD range of 2.77–6.77), as well as using the uncertainty estimates on the serial interval mean and SD reported in Reed et al.(12), neither of which substantially changed the estimates for R_t_ or when R_t_ crossed 1 (not shown).

### Human Subjects Approval

Sequencing of samples was approved by the University of Michigan Institutional Review Board (IRB Protocol ID: HUM185966) or performed as part of a public health investigation prompted by state and local health officials (IRB Protocol ID: HUM198210).

## Results

Twenty-six first-level contacts (close contacts and team members) were identified. Ten of these first-level contacts subsequently became positive for SARS-CoV-2. An additional 4 cases were identified among 38 contacts of contacts (i.e. second-level contacts). In total, 15 cases with an epidemiologic linkage to cluster A were identified over 18 days. During the investigation of the initial cluster, seven of the 10 initial close contacts with SARS-CoV-2 were confirmed as lineage B.1.1.7, and one of the 4 second-level contacts with SARS-CoV-2 was confirmed as B.1.1.7 (Figure 1, Cluster A). The remaining six cases among first and second-level contacts did not have residual specimens available for characterization.

**Figure 1.**
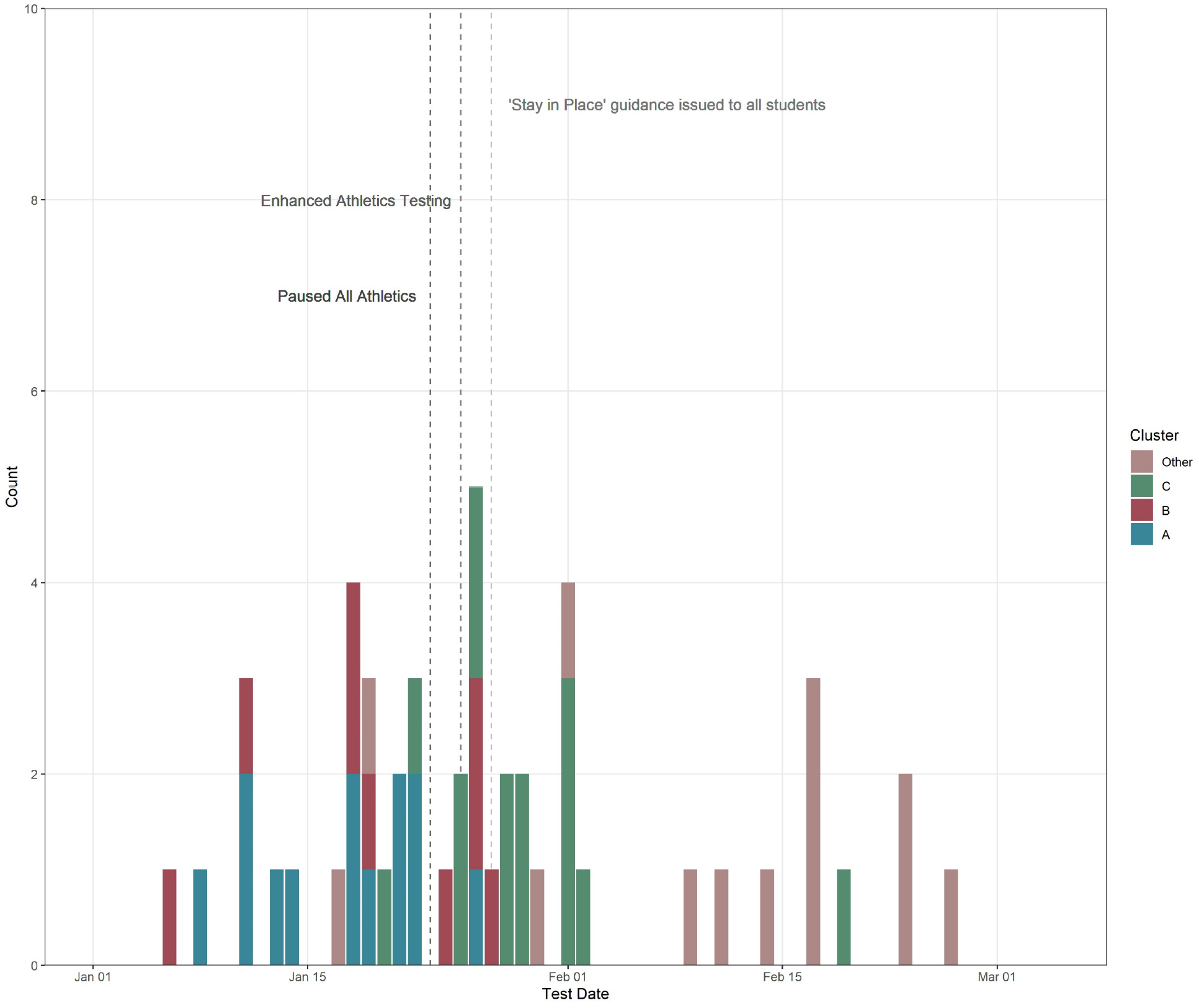
Epidemiologic curve of B.1.1.7 cases, by cluster Count of cases by cluster (Groups A, B, and C) are plotted by the earliest of the date of symptom onset, when available, or date of positive test. B.1.1.7 cases not linked to one of the three cluster groups are listed as “Other”. Dashed lines indicate dates of mitigation measures.

Expanded surveillance efforts identified a second cluster of B.1.1.7 with epidemiologic linkage to an individual with a test collection date that preceded cluster A, a history of domestic travel, and participation on an unrelated athletic team (Figure 1, Cluster B). For the cluster B index case, 43 close contacts were identified, of whom 14 became cases; four of which were identified by sequencing as having B.1.1.7. Among 17 second-level contacts, 5 additional cases were identified, but none had specimens available for sequencing.

In late January, a third travel-associated cluster was identified in a separate group of students with no connection to clusters A or B (Figure 1, Cluster C). This cluster had a total of 24 epidemiologically-linked cases; 12 of which were confirmed to be B.1.1.7. Similarly to clusters A and B, infections were identified in first and second level contacts, but further spread was not identified. Notably, weekly asymptomatic testing was continued for all students on campus, and increased in frequency for student athletes, throughout the investigation of the three clusters.

Viral sequences were used to determine that the three initial B.1.1.7 clusters were distinct and unconnected. Genomes in all three clusters were identified as B.1.1.7 and contained the 17 lineage-defining mutations. Virus genomes across the three clusters were genetically distinct, separated by inter-cluster distances of five single nucleotide substitutions or greater. Mutations C12068T, A17615G, and C25613T were unique to genomes in cluster A; mutations C2110T, G3004T, G7042T, C14120T, and A28095T were unique to genomes in cluster B; mutations C5192T and C27513T were unique to genomes in cluster C.

Cases and contacts were initially interviewed at the time of SARS-CoV-2 detection following standard county health department protocols, and all confirmed and probable B.1.1.7 cases were subsequently re-interviewed by case investigators. Although a number of positive cases were identified through initial asymptomatic testing, the majority of cases of both B.1.1.7 and non-B.1.1.7 cases identified from January 6 through February 26 were symptomatic at the time of case interview (95% symptomatic for B.1.1.7 versus 89% for non-B.1.1.7; p=0.15 by Mid-P exact test).

Attack rate was calculated for first-level contacts and second-level contacts, excluding contacts who had previous infections within the last 90 days. Within clusters A and B, which resulted from independent introductions, inclusive of susceptible contacts and second-level contacts, the secondary attack rate for confirmed B.1.1.7 cases was 25.8% (32 of 124; (Wilson 95% Confidence Interval: 0.20, 0.34). R_t_ was estimated for all B.1.1.7 cases on campus beginning in January through late February. R_t_ initially increased in mid-January, followed by a decline to below 1 as the three initial clusters resolved in late January and early February. R_t_ increased again after the second week of February with the occurrence of additional B.1.1.7 introductions. (Figure 2)

**Figure 2.**
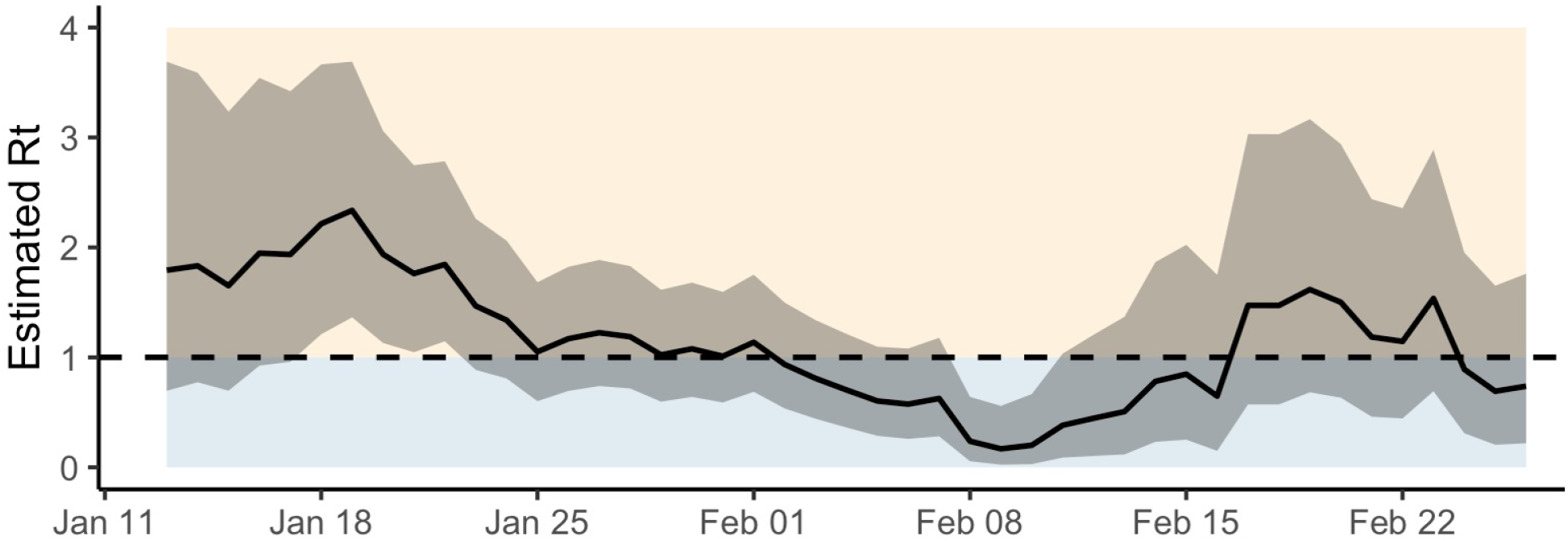
Estimated R_t_ for UM B.1.1.7 cases. Median R_t_ is plotted by time (x-axis). Grey shaded region is the 95% confidence interval. R_t_ = 1 is indicated by dashed line.

The larger Washtenaw County community was sampled during the period of January 1, 2021 through February 28, 2021 by inclusion of all available positive specimens from Michigan Medicine, a large academic medical center. Sequencing revealed a diverse outbreak with multiple circulating lineages both on campus and across the larger Washtenaw County community. While B.1.1.7 lineage viruses circulated in both the campus population and the non-campus community, the three clusters identified were not connected to further non-campus spread (Figure 3). Additional introductions and transmission events beyond the three initial clusters were identified beginning in late February.

**Figure 3.**
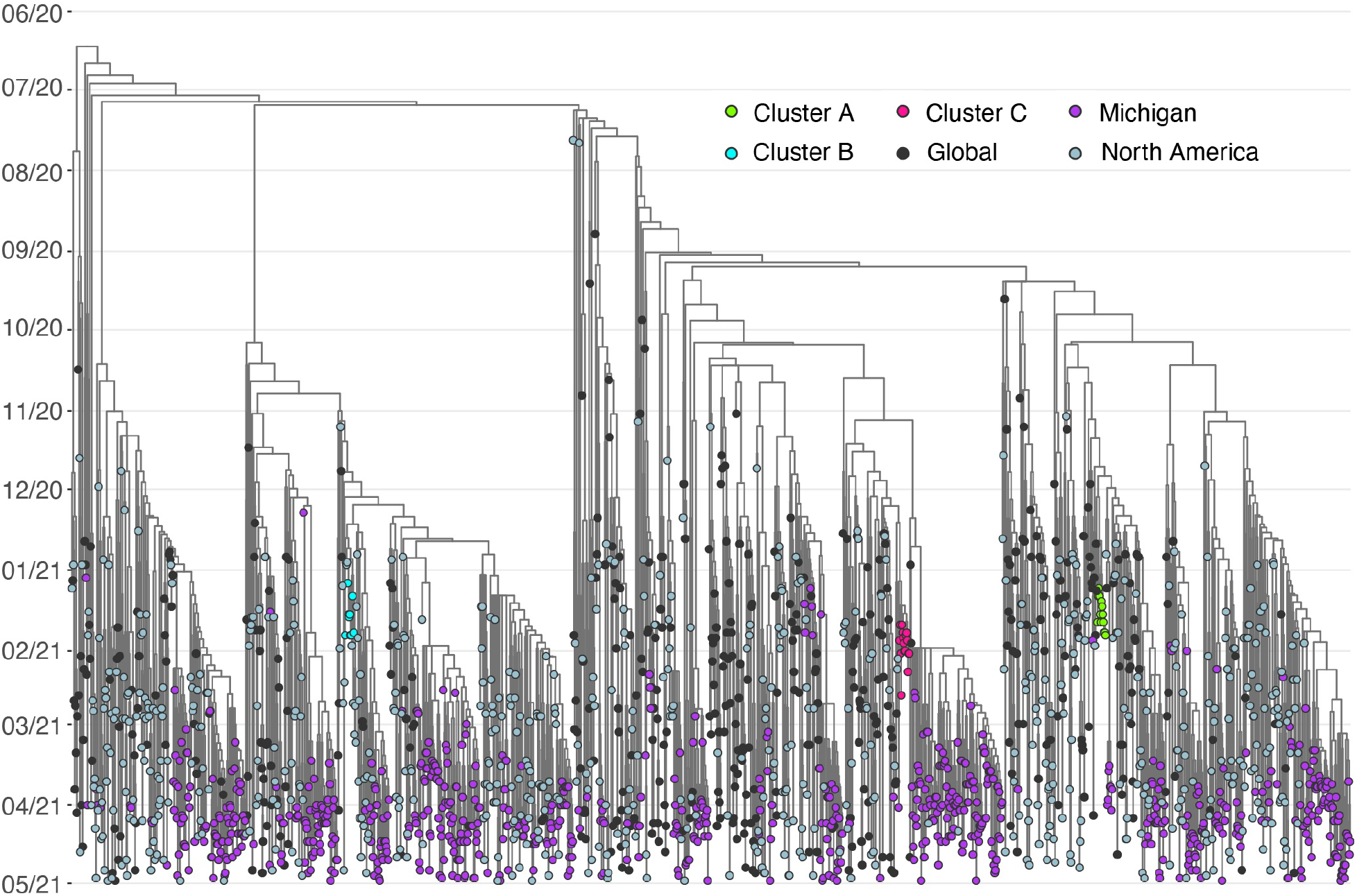
Phylogenetic tree of three B.1.1.7 clusters alongside regional, national, and global B.1.1.7 sequences. Time-calibrated maximum likelihood phylogenetic tree of 1958 B.1.1.7 genomes, including 804 B.1.1.7 genomes sequenced in this study. Month is shown on the x-axis. Tips are colored as follows: Cluster A (green), Cluster B (cyan); Cluster C (red); B.1.1.7 cases presenting for testing to University Health Services, the university asymptomatic surveillance testing program, or Michigan Medicine (purple); global (black) and North American (grey) contextual genomes. No descendants of Clusters A-C were identified.

### Public Health Response

The university collaborated with local and state health departments to rapidly implement multiple strategies targeted at containment of B.1.1.7 transmission. Antigen testing protocols were temporarily supplemented with additional PCR-testing three times per week for all athletic teams participating in conference mandated rapid antigen testing protocols. All athletics competitions and practices were paused per MDHHS guidance from January 23 to February 7, 2021. Quarantine criteria was extended to include all athletic team members including those not specifically identified as close contacts. Students traveling to the area were asked to test within 72 hours of arrival on campus, and ongoing weekly PCR tests were required for all students throughout the semester(14). Investigation and mitigation of the two B.1.1.7 clusters occurred at the same time as a large, coincident increase in non-B.1.1.7 transmission in the campus community. On January 27, 2021, a school-wide stay-at-home recommendation was issued for the general university student population through February 7. Additional testing was offered by the health department to the local non-university community in major locations near campus. Public notifications were released identifying possible times and locations of potential community exposures to B.1.1.7. No additional cases linked to these exposures were identified.

## Discussion

The worldwide emergence of a succession of variants of concern, and the resulting impacts on disease incidence, are a stark demonstration of the critical importance of continued genomic tracking throughout the next phase of the COVID-19 pandemic. The outbreak described here provided a unique opportunity to directly observe lineage-specific emergence and spread and to initiate genomically-informed public health response at the local level. The processes used here are particularly informative as universities and other communities plan responses to Omicron, which also can be detected through an SGTF pattern, enabling efficient use of RT-PCR screening followed by targeted sequencing.

Here we report the early identification of three initial introductions of an emerging lineage into a college campus, a high-risk population for SARS-CoV-2 spread(5,15,16) and emergence of travel-associated variants(4). The investigation benefited from available infrastructure for widescale asymptomatic testing and for genomic surveillance. This infrastructure substantially increased the speed with which the first B.1.1.7 case was identified. By virtue of extensive sequencing and weekly testing of the campus population in January and February 2021 (over 1,300 specimens sequenced in January and February), we were able to document three introduction events, all resulting from travel. There are a number of lessons learned from this experience that can directly inform the response of the international community to additional VOCs. We used just-in-time sequencing to deploy a genomically-informed mitigation strategy that included enhanced case investigation and contact tracing methods and frequent testing in the specific communities with exposure to B.1.1.7. We paired this strategy with initial testing with a RT-PCR assay to distinguish non-B.1.1.7 cases, enabling us to prioritize sequencing resources accordingly. These methods were ultimately successful in preventing larger community transmission resulting from the first two transmission events, thus delaying widespread predominance of B.1.1.7 for multiple weeks. Importantly, genomic data made it clear when the mitigation measures were successful for specific B.1.1.7 clusters. Clusters ceased to have genomic descendants once transmission was controlled, which allowed measures targeted towards specific risk cohorts to be ended.

The investigation described here has shown clearly that the emergence of B.1.1.7 into the community was not due to a single introduction. This is most apparent on this smaller scale, where individual clusters can be characterized, and this is consistent with patterns of emergence that are observed at a larger level, through counties, states and communities. Simplistic epidemiologic curves of cases give the impression that a single initiating infection led to an exponential rise. However, the addition of genomic data, such as that presented here, makes it clear that introductions are complex and multifaceted, with what appeared to be a single outbreak due to 3 introductions. Genomic data also demonstrated little genetic similarity between these initial clusters and viruses that ultimately emerged in the broader community, a pattern noted previously in our area (17).

While a number of both B.1.1.7 and non-B.1.1.7 cases during this time period were first identified through asymptomatic surveillance, it is notable that almost all cases eventually reported symptoms at some point during the case investigation and follow-up process, with 95% of B.1.1.7 cases reporting symptoms. This finding is in contrast to other evaluations that have reported the fraction of completely asymptomatic cases to be above 25%(18–20). This difference could be explained by population differences, or differences in intensity of follow-up for symptoms in the context of an investigation of an emerging variant. Our data underscore the fact that the asymptomatic proportion of a COVID-19 outbreak may be overestimated, and thus the clinical impact underestimated, when subsequent development of symptoms is not ascertained in the days following testing(21,22).

As SARS-CoV-2 variants continue to emerge, these results demonstrate that high-resolution testing and sequencing, when coupled with targeted public health mitigation measures (increased frequency of testing, expanded case investigation and contact tracing interviews, and enhanced quarantine practices), can delay larger dissemination of a more transmissible strain in a campus setting. This strategy can inform efforts to control future SARS-CoV-2 variant emergence as colleges and communities target genomic surveillance resources into the next phase of the pandemic.

## Experimental Procedures

Residual transport media or saliva was centrifuged at 1200 x g. and aliquoted. For nasopharyngeal and sputum specimens, RNA was extracted with the Invitrogen PureLink Pro 96 Viral RNA/DNA Purification Kit (200 μL of input sample eluted in 100 µL) or the QIAamp Viral RNA Mini kit (140 µL of input sample eluted in 50 µL). For saliva specimens, RNA was extracted with the Thermo Fisher MagMAX Viral RNA Isolation Kit (200 µL of input sample eluted in 50 µL). Extracted RNA was reverse transcribed with SuperScript IV (Thermo Fisher). For each sample, 1 µL of random hexamers and 1 µL of 10 mM dNTP were added to 11 µL of RNA, heated at 65°C for 5 min, and placed on ice for 1 min. Then a reverse transcription master mix was added (4 µL of SuperScript IV buffer, 1 µL of 0.1M DTT, 1 µL of RNaseOUT RNase inhibitor, and 1 µL of SSIV reverse transcriptase) and incubated at 42 °C for 50 min, 70 °C for 10 min, and held at 4 °C. SARS-CoV-2 cDNA was amplified in two multiplex PCR reactions with the ARTIC Network version 3 primer pools and protocol. Viral cDNA was amplified with the Q5 Hot Start High-Fidelity DNA Polymerase (NEB) with the following thermocycler protocol: 98 °C for 30 s, then 35 cycles of 98 °C for 15 s, 63 °C for 5 min, and final hold at 4 °C. Reaction products for a given sample were pooled together in equal volumes.

### Illumina library preparation and sequencing

Pooled PCR product was purified with 1X volume of AMPure beads (Beckman-Coulter). Sequencing libraries were prepared with the NEBNext Ultra II DNA Library Prep Kit (NEB) according to the manufacturer’s protocol. Barcoded libraries were pooled in equal volume and extracted with a 1% agarose gel to remove adapter dimers. Pooled libraries were quantified with the Qubit 1X dsDNA HS Assay Kit (Thermo Fisher). Libraries were sequenced on an Illumina MiSeq (v2 chemistry, 2×250 cycles) at the University of Michigan Microbiome Core facility. Reads were aligned to the Wuhan-Hu-1 reference genome (GenBank MN908947.3) with BWA-MEM version 0.7.15. Sequencing adaptors and amplification primer sequences were trimmed with iVar 1.2.1. Consensus sequences were called with iVar 1.2.1 by simple majority at each position (>50% frequency), placing an ambiguous N at positions with fewer than 10 reads.

### Oxford Nanopore library preparation and sequencing

After multiplex PCR amplification, libraries were prepared for sequencing with the Oxford Nanopore Technologies MinION using the ARTIC Network version 3 protocol (6,7). Samples were prepared in batches of 24 with one-pot native barcoding. Pooled PCR products were diluted in nuclease-free water with a dilution factor of 10. Amplicon ends were prepared for ligation with the NEBNext Ultra II End Repair/dA-Tailing Module (NEB). Unique barcodes (Oxford Nanopore Native Barcoding Expansion kits) were ligated per sample with the NEB Blunt/TA Ligase Master Mix. After barcoding, reactions were pooled together in equal volumes and purified barcoded amplicons with 0.4X volume of AMPure beads. Oxford Nanopore sequencing adapters were ligated with the NEBNext Quick Ligation Module (NEB) and the library was purified with 1X volume of AMPure beads. Final libraries were quantified with the Qubit 1X dsDNA HS Assay Kit (Thermo Fisher). Each library (15-20 ng) was loaded onto a flow cell (FLO-MIN106) and sequenced with the MinION.

## Supporting information

Strobe Checklist

## Data Availability

All data produced in the present study are available upon reasonable request to the authors if approved by the Insitutional Review Boards of participating insitutions.

## Acknowledgments

Kalee Rumfelt, Chris Blair, Will Fitzsimmons, Jennifer Nord, Christine Kizer, Jennifer Bergendahl, University of Michigan; Laura Bauman, Washtenaw County Health Department; Diana Riner, PhD, Michigan Department of Health and Human Services; Microbiology, Virology, and Bioinformatics sections, Michigan Department of Health and Human Services. Sequencing was supported by CDC contract 75D30120C09870 (to ASL) and University COVID response funds.

---

**Supplementary Figure.**
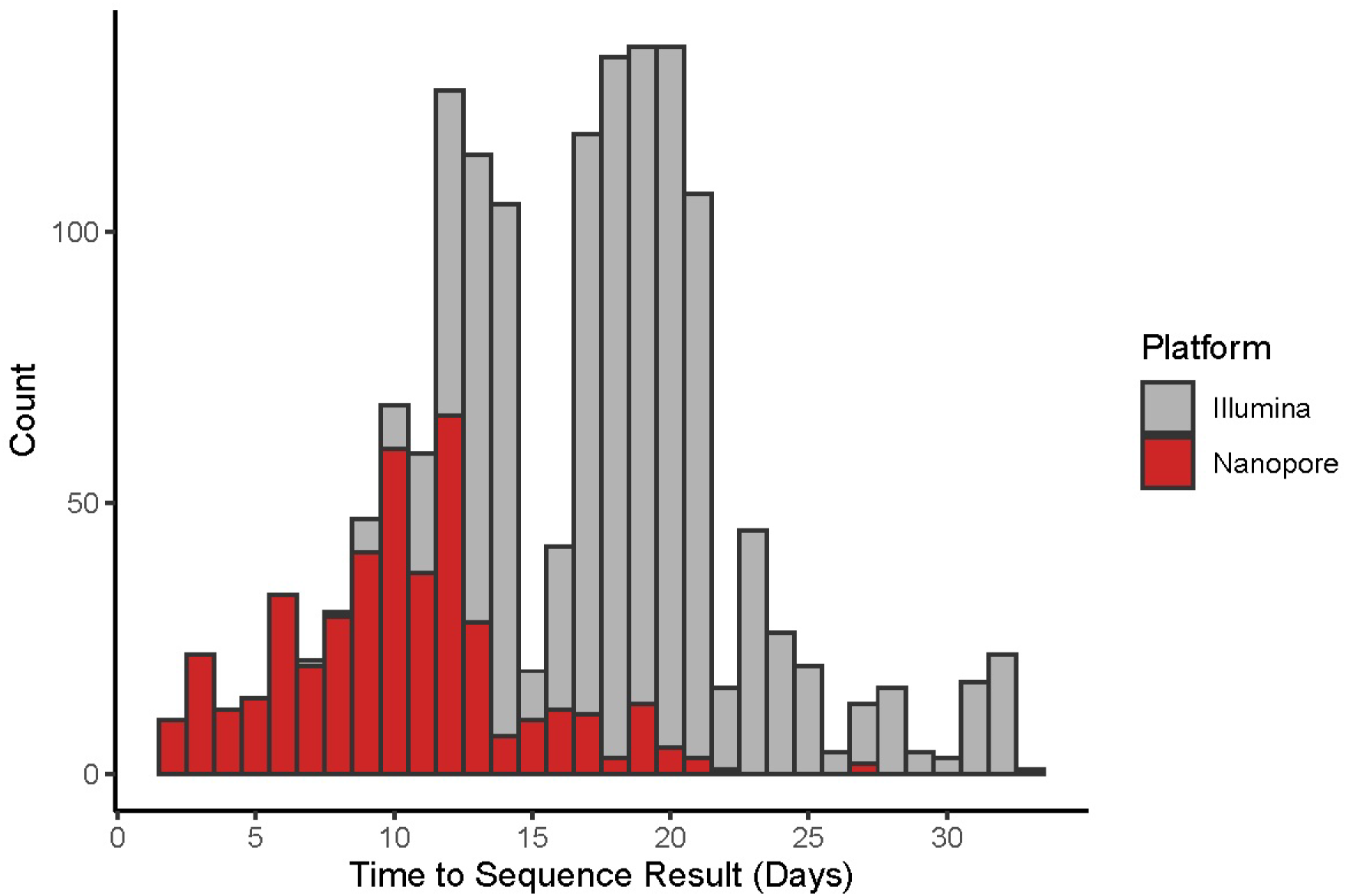
Time between positive result and availability of sequence result. Specimen count is plotted by turnaround time (number of days between positive result and completion of sequencing). Bars indicate specimens tested by Nanopore (red) and Illumina (gray) platforms.

## Notes

### Competing Interest Statement

All authors have completed the ICMJE uniform disclosure form at www.icmje.org/coi_disclosure.pdf and declare: Financial support for the submitted work from CDC and from university COVID response funds. E.T.M. has received grant funding from CDC, NIH, and Merck. A.S.L. has received grant funding from CDC, NIH, and Burroughs Wellcome Fund and consulting fees from Sanofi and Roche. Y.N. is co-owner of LynxDx, Inc. D.C. has received speaker fees from National Athletic Trainers' Association, Far West Athletic Trainers' Association, and Tenessee Athletic Trainers' Society and consulting fees from The Rehberg-Konin Group

### Author Declarations

University of Michigan Institutional Review Board

